# Relationships between barley consumption and gut microbiome characteristics in a healthy Japanese population: a cross-sectional study

**DOI:** 10.1101/2021.09.13.21263482

**Authors:** Tsubasa Matsuoka, Koji Hosomi, Jonguk Park, Yuka Goto, Mao Nishimura, Satoko Nakashima, Haruka Murakami, Kana Konishi, Motohiko Miyachi, Hitoshi Kawashima, Kenji Mizuguchi, Toshiki Kobayashi, Hiroshi Yokomichi, Jun Kunisawa, Zentaro Yamagata

**Affiliations:** Research and Development Department, Hakubaku Co., Ltd., 4629, Nishihanawa, Chuo, Yamanashi 409-3822, Japan; Department of Health Sciences, School of Medicine, University of Yamanashi, 1110, Shimokato, Chuo, Yamanashi 409-3898, Japan; Laboratory of Vaccine Materials, Center for Vaccine and Adjuvant Research, National Institutes of Biomedical Innovation, Health and Nutrition, 7-6-8 Asagi, Saito, Ibaraki, Osaka 567-0085, Japan; Laboratory of Bioinformatics, National Institutes of Biomedical Innovation, National Institutes of Biomedical Innovation, Health and Nutrition, 7-6-8 Asagi, Saito, Ibaraki, Osaka 567-0085, Japan; Department of Physical Activity Research, National Institutes of Biomedical Innovation, Health and Nutrition, 1-23-1 Toyama, Shinjuku, Tokyo 162-8636, Japan; Institute for Protein Research, Osaka University, Osaka, Japan; Department of Microbiology and Immunology, Kobe University, Hyogo, Japan; Graduate Schools of Medicine, Pharmaceutical Sciences, and Dentistry, Osaka University, Osaka, Japan; Department of Microbiology and Immunology and International Research and Development Center for Mucosal Vaccines, The University of Tokyo, Tokyo, Japan; Faculty of Science and Engineering, Waseda University, Tokyo, Japan

**Keywords:** barley, beta-glucan, microbiome, *Bifidobacterium*, *Anaerostipes*, *Butyricicoccus*

## Abstract

**Objective:** Barley contains abundant soluble β-glucan fibres, which have established health benefits. In addition, the health benefits conferred by the gut microbiota have attracted considerable interest. However, few studies have focused on the barley intake and microbiota of the Japanese population. In this study, we aimed to identify the relationship between the barley consumption and gut microbiota composition of the Japanese population.

**Research Methods & Procedures:** A total of 236 participants were recruited in Japan, and 94 participants with no complication of diabetes, hypertension, and dyslipidemia were selected for the study. We analysed faecal samples from the participants, their medical check-up results, and responses to questionnaires about dietary habits. The participants were grouped according to their median barley intake. Then, we assessed the relative abundance of 50 microbial genera throughout the group and selected 20 that differed at *P <* 0.1 (Mann–Whitney *U*-test). We also analysed the networks and clustering of the 20 selected genera.

**Results:** According to their relative abundance, *Bifidobacterium, Anaerostipes*, and *Butyricicoccus* were candidate characteristic microbiota of the group that consumed large amounts of barley (*P* < 0.05). Furthermore, network and cluster analyses revealed that barley directly correlated with *Bifidobacterium* and *Butyricicoccus*.

**Conclusions:** Barley consumption generates changes in the intestinal microbiota of the Japanese population. We found that *Bifidobacterium, Anaerostipes*, and *Butyricicoccus* abundance was positively associated with barley consumption. This study showed that barley is important for gut microbiota and relates to Japanese traditional food like natto.

## Introduction

The gut microbiota is important for health, and thus, several investigations have focused on the human gut microbiome. It has been reported that diet can alter the gut microbiome ^(1)^. Microbiota-accessible carbohydrates, such as dietary fibre ^(2)^, are resistant to digestion in the small intestine and enter the large intestine undigested; therefore, they are likely to improve host metabolism ^(2)^.

Barley is an important cereal that contains the soluble fibre β-glucan ^(3)^. Barley improves metabolic dysfunction, increases the diversity of the gut microbiota, and increases bacteria such as *Blautia* ^(4)^. Additionally, barley lowers postprandial blood glucose in healthy ^(3)^ and diabetic patients ^(5)^, and lowers cholesterol concentrations in Japanese people with mild metabolic syndrome after 12 weeks of consumption ^(6)^. Therefore, barley consumption has many potential benefits to global health.

Barley consumption affects the gut microbiota and host health ^(4, 7, 8)^. For example, a crossover study in Sweden found that barley intake increased blood concentrations of butyric acid (produced by intestinal bacteria) and decreased postprandial hyperglycaemia ^(7)^. Research in the USA found that barley intake increased the gut microbiota diversity and *Blautia* abundance and improved host cholesterol levels ^(4)^. Another Swedish study found that barley intake promoted a high ratio of *Prevotella*/*Bacteroide*s and improved host blood glucose metabolism ^(4)^. These results suggest that barley might beneficially modulate the composition of the gut microbiota and improve host metabolic health.

Indeed, the effects of barley on the gut microbiota have been established in several countries ^(4, 7, 8)^. Furthermore, as the intestinal microbiota is affected by dietary habits and genetic factors, it is important to evaluate its effectiveness in different populations ^(9)^. The Japanese gut microbiota is characterised by abundant *Blautia* and *Bifidobacterium*, which utilise the dietary fibre from seaweeds, a common item in the Japanese diet ^(1)^. Consuming barley with white rice is also a specific dietary habit in Japan, but little is known about its effects on the microbiota. Most studies on barley and the microbiota have regarded exhaled hydrogen concentrations as an alternative variable of intestinal bacteria ^(10, 11, 7)^ ; however, few studies have analysed the relationship between the microbiota and barley consumption. Therefore, we aimed to define the relationship between barley and gut microbiota composition in healthy Japanese adults using next-generation sequencing.

## Methods

### Study design

The study was approved by the Yamanashi University Ethics committee (approval No.1824), National Institutes of Biomedical Innovation, Health and Nutrition Ethics committee (approval No. 169-04), and Chiyoda Paramedical Care Clinic Ethics committee (approval No. 15000088). This study was conducted in accordance with the Declaration of Helsinki (2013) and was based on a registered study (UMIN000033479). Cross-sectional data were evaluated from the first year of the study to undertake an exploratory overview of the gut microbiome of a population that consumes barley. Sampling was conducted from August 2018 to March 2019.

We enrolled 272 participants, which were employees of the barley processing company Hakubaku Co., Ltd. Our target sample included at least 100 participants. We excluded those with disorders (Risk 2) and pre-disorders (Risk 1) of diabetes, hypertension, and dyslipidemia from the main analysis. Details of the exclusion criteria for disorders are shown in Supplemental Table 1. We classified the participants into two groups based on the median barley consumption rate (high, 32.3–253 and low, 0–31.8 g/1,000 kcal·day).

### Measurements

The primary outcome was the association between barley consumption and the alpha-diversity of the microbiome, and the secondary outcome was the abundance of the 50 dominant microbiota sorted by mean relative abundance (Supplemental Tables 3 and 4). We collected a copy of the participant’s medical check-up results. Dietary habits other than barley consumption were assessed using a brief self-administered diet history questionnaire (BDHQ) (Gender Medical Research, Inc., Tokyo, Japan). Barley consumption (g/1,000 kcal·day) was calculated using a questionnaire and the daily energy value from the BDHQ. Rice bowl size (200, 160, 140, and 100 g), proportion of barley mixed with white rice (0%, 5%, 10%, 15%, 30%, and 50%), barley-mixed rice consumed per month (0, 0.5, 1, 4, 8, and 16 days/month), and barley consumption rate (g/day) were determined. Medical history, including medication (especially during the month of sampling), and consumption of fermented foods and supplements were determined using questionnaires.

### DNA extraction and 16S rRNA gene amplicon sequencing

Faecal samples were collected at home with guanidine thiocyanate (GuSCN) solution; and DNA was extracted and stored at 10–30°C within 30 days until DNA extraction ^(12)^. Briefly, 0.2 mL of faecal samples, 0.3 mL of No. 10 lysis buffer (Kurabo Industries Ltd., Osaka, Japan), and 0.5 g of 0.1-mm glass beads (WakenBtech Co., Ltd., Tokyo, Japan) were homogenised using a PS1000 Cell Destroyer (Bio Medical Science, Tokyo, Japan) at 4,260 rpm for 50 s at 25°C. The homogenate was centrifuged at 13,000 × g for 5 min at 25°C, and the DNA was extracted from the supernatant using a Gene Prep Star PI-80X automated DNA isolation system (Kurabo Industries Ltd). DNA concentration was determined with the ND-1000 NanoDrop Spectrophotometer (Thermo Fisher Scientific Inc., Waltham, MA, USA). The samples were stored at -30°C. The 16S rRNA gene was amplified from faecal DNA and sequenced ^(12)^. The V3–V4 region of the 16S rRNA gene was amplified using the following primers (5′→3′): TCGTCGGCAGCGTCAGATGTGTATAAGCGACAGCCTACGGGNGGCWGCAG and GTCTCGTGGGCTCGGAGATGTGTATAAGAGACAGGACTACHVGGGTATCTAATCC. The DNA library for Illumina MiSeq was prepared using Nextera XT Index Kit v2 Set A (Illumina Inc., San Diego, CA, USA), and its concentration was determined with the QuantiFluor dsDNA System (Promega Corp., Madison, WI, USA). The 16S rRNA gene was sequenced using Illumina MiSeq (Illumina) as described by the manufacturer.

### Bioinformatics analysis

The sequence reads from Illumina MiSeq were analysed using the Quantitative Insights Into Microbial Ecology (QIIME) software package (v1.9.1) ^(13)^. We used QIIME Analysis Automating Script (Auto-q) ^(14)^ to proceed from trimming paired-end reads to operational taxonomic unit (OUT) selection. We used open-reference OTU picked with the UCLUST software against the SILVA v128 reference sequence to select OTUs based on sequence similarity (>97%). The taxonomy (phylum, class, order, family, and genus) and relative abundance were calculated using the SILVA v128 database ^(13, 14)^. The intestinal microbiota was compared in 10,000 randomly selected reads per sample.

### Statistical analyses

#### Calculation of alpha-diversity

Data were exported as BIOM files and imported into R (version 3.6.0). Diversity was analysed using the phyloseq R-package. Alpha-diversity indices of observed OTU, Chao 1, Shannon, and Simpson indices were calculated using the estimate_richness function.

#### Comparison of barley groups

The results of the medical check-ups and dietary habits between the high and low barley groups were compared using Student’s *t*-tests. The alpha-diversity and relative abundance of each genus were analysed using Mann–Whitney *U*-tests. *P* values were adjusted using false discovery rate (FDR) methods. To confirm the reliability of the analyses, we explored the relationship between barley consumption rates and each microbiome using multiple regression analyses with the groups (0 = low and 1 = high) or consumption rate (g/1,000 kcal·day) and Kendall rank-sum tests. We set the amounts of *Butyricicoccus, Bifidobacterium, Ruminococcus 2, Anaerostipes*, and the barley groups (0 = low and 1 = high) as independent variables in the multiple regression analyses. We adjusted the model for age, sex, and the consumption rate of dietary factors (g/1,000 kcal) as independent variables. Correlations between barley and dietary habits were evaluated using Kendall rank-sum tests referring to published criteria ^(17)^.

#### Principal coordinate analysis of microbiomes

We classified the participants into enterotypes A, B, and C using the pam function of the cluster R-package. We then summarised the composition of intestinal microbiota by principal coordinate analysis (PCoA) using the vegdist function of the vegan R-package and the quasieuclid and dudi.pco functions of the ade4 R-package. Data were calculated using the Bray–Curtis method. Subsequently, the environmental factor arrows were fit to the PCoA figure using the envfit function of the vegan R-package. Significant genera were assessed using permutations of environmental variables.

#### Network analysis of significant microbiomes and barley

To visualise the associations between barley and 20 microbiome genera selected by *P* < 0.1, we implemented a network analysis. The network is shown with lines of correlation with |*r*| > 0.15 (Kendall rank-sum tests). Different colours on the plots indicate different community groups. We fit the correlation data frame to the cc.df function of the igraph R-package using the reshape2 R-package and calculated microbiota community groups using the leading.eigenvector.community function.

All analyses were carried out in R (version 3.6.0), and tests were two-sided; *P* < 0.05 was considered significant. All graphs except for those from the network analysis were created with the R-package ggplot2 ^(18)^.

#### Effects of barley intake on the participants

Associations between barley and *Butyricicoccus, Bifidobacterium, Ruminococcus 2*, and *Anaerostipes* were assessed in all participants using multiple regression analyses. We set each genus as the dependent variable and barley groups (0 = low, 1 = high), sex, age, risk of diabetes, dyslipidemia, and hypertension (0–2) as the independent variables. We used the vif function of the car R-package to evaluate variance inflation factors (VIFs). All VIFs were < 5 and considered acceptable for these analyses^(19)^.

## Results

### Participants

We obtained informed consent from 272 individuals, of which 33 participants resigned and 6 were excluded because of non-compliance and data loss. Ninety-four participants had no disorders and were included in the main analyses and 236 were included in the multiple regression analyses (Figure 1).

**Figure 1.**
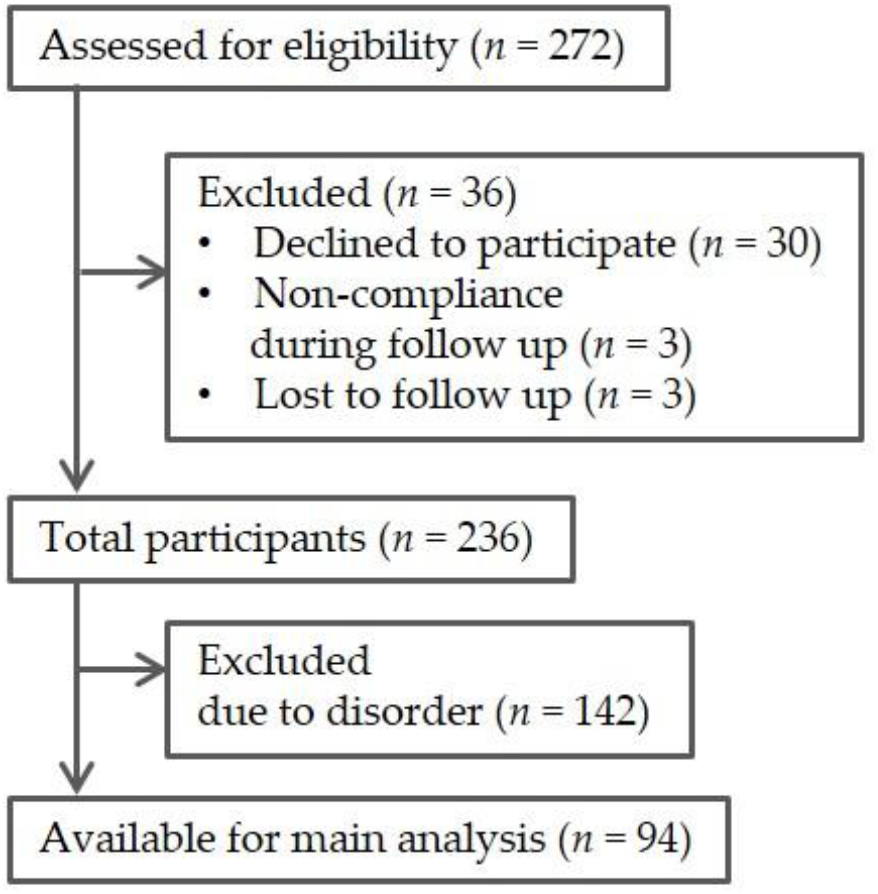
Flow chart of participants. Disorder means risk of diabetes, hypertension, and dyslipidemia.

The characteristics of the barley groups did not significantly differ (Table 1). However, within the normal range, fasting glucose concentrations tended to be higher in the high barley group (*P* = 0.054). Disorder risk did not significantly differ between the groups (**Supplemental Table 1**). **Table 2** shows the differences in dietary habits between the barley groups. The consumption rates of dietary fibre (*P* < 0.001), cereal (*P* = 0.048), and natto (soybeans fermented with *Bacillus subtilis*; *P* < 0.001) were significantly higher in the high barley group and those of legumes (*P* = 0.06) tended to be higher in the high barley group. Therefore, we selected these four dietary factors for adjustment in the multiple regression analyses.

**Table 1.** Characteristics of the participants in the barley groups

| Variable | Total ($n = 94$) | Low barley<sup>1</sup> | High barley<sup>1</sup> | $P^2$ |
| --- | --- | --- | --- | --- |
|  | 0–253 | 0–31.8 | 32.3–253 |  |
|  | g/1,000 kcal d | g/1,000 kcal·d | g/1,000 kcal·d |  |
| Male, $n$ (%) | 54 (57%) | 32 (68%) | 22 (47%) | |
| Age (years) | $36 \pm 10$ | $36 \pm 10$ | $36 \pm 11$ | 0.82 |
| Body mass index ($\text{kg}/\text{m}^2$) | $21.5 \pm 3.4$ | $21.3 \pm 4.1$ | $21.6 \pm 2.5$ | 0.67 |
| Systolic pressure (mmHg) | $111 \pm 11$ | $110 \pm 11$ | $112 \pm 11$ | 0.25 |
| Diastolic pressure (mmHg) | $68 \pm 9$ | $67 \pm 9$ | $69 \pm 8$ | 0.39 |
| Fasting glucose (mg/dL) | $86 \pm 7$ | $84 \pm 7$ | $87 \pm 7$ | 0.054 |
| HbA1c (%) | $5.3 \pm 0.2$ | $5.3 \pm 0.2$ | $5.3 \pm 0.2$ | 0.26 |
| TG (mg/dL) | $69 \pm 28$ | $67 \pm 30$ | $71 \pm 26$ | 0.42 |
| HDL-cholesterol (mg/dL) | $65 \pm 13$ | $66 \pm 13$ | $63 \pm 12$ | 0.26 |
| LDL-cholesterol (mg/dL) | $95 \pm 15$ | $93 \pm 14$ | $97 \pm 16$ | 0.15 |
| *Daily Nutritional intake* |  |  |  |  |
| Energy (kcal) | $1729 \pm 510$ | $1693 \pm 456$ | $1765 \pm 562$ | 0.50 |
| Protein (g/1,000 kcal) | $34 \pm 6$ | $35 \pm 6$ | $34 \pm 6$ | 0.58 |
| Fat (g/1,000 kcal) | 30 ± 6 | 30 ± 6 | 30 ± 6 | 0.85 |
| Carbohydrate (g/1,000 kcal) | 130 ± 19 | 130 ± 20 | 129 ± 18 | 0.75 |
| Na (mg) | 3740 ± 1027 | 3608 ± 948 | 3871 ± 1094 | 0.22 |

Data are shown as means ± SD or n (%). <sup>1</sup>Range of barley consumption.
<sup>2</sup>Student's *t*-test

**Table 2.** Comparison of diet between the barley groups

| Variable | Low barley<sup>1</sup> (*n* = 47)<br>0–31.8 g/1,000 kcal·d | High barley<sup>1</sup> (*n* = 47)<br>32.3–253 g/1,000 kcal·d | *P*<sup>2</sup> |
| --- | --- | --- | --- |
| Barley (g/1,000 kcal) | 13.9 ± 11.3 | 70.9 ± 43.6 |  |
| Beta-glucan (g/1,000 kcal) | 0.70 ± 0.57 | 3.6 ± 2.2 |  |
| Carbohydrate (g/1,000 kcal) | 129 ± 18 | 130 ± 20 | 0.75 |
| Dietary fibre (g/1,000 kcal) | 4.9 ± 1.3 | 6.1 ± 1.7 | <0.001 |
| Cereal (g/1,000 kcal) | 213 ± 55 | 236 ± 58 | 0.048 |
| Legumes (g/1,000 kcal) | 20 ± 15 | 27 ± 20 | 0.06 |
| Green vegetable (g/1,000 kcal) | 37 ± 23 | 44 ± 32 | 0.25 |
| Other vegetable (g/1,000 kcal) | 62 ± 29 | 72 ± 45 | 0.20 |
| Fruit (g/1,000 kcal) | 36 ± 32 | 35 ± 30 | 0.87 |
| Seaweed (g/1,000 kcal) | 3.5 ± 2.8 | 4.0 ± 3.9 | 0.51 |
| Natto (g/1,000 kcal) | 3.6 ± 4.2 | 10.4 ± 12 | <0.001 |
| Mushroom (g/1,000 kcal) | 4.4 ± 2.5 | 5.0 ± 4.1 | 0.37 |

Data are shown as means ± SD. <sup>1</sup>Range of barley consumption.
<sup>2</sup> Student's *t*-test

### Enterotypes

**Figure 2a** describes the number of participants with each enterotype. Enterotype B was dominant and enterotypes A and C were less abundant in each group. The numbers of each enterotype did not significantly differ between the groups (*P =* 0.20). Figure 2b shows an overview of the PCoA of the microbiome. The distribution of the plots was laid out in a “∧” shape and separated into three groups.

**Figure 2.**
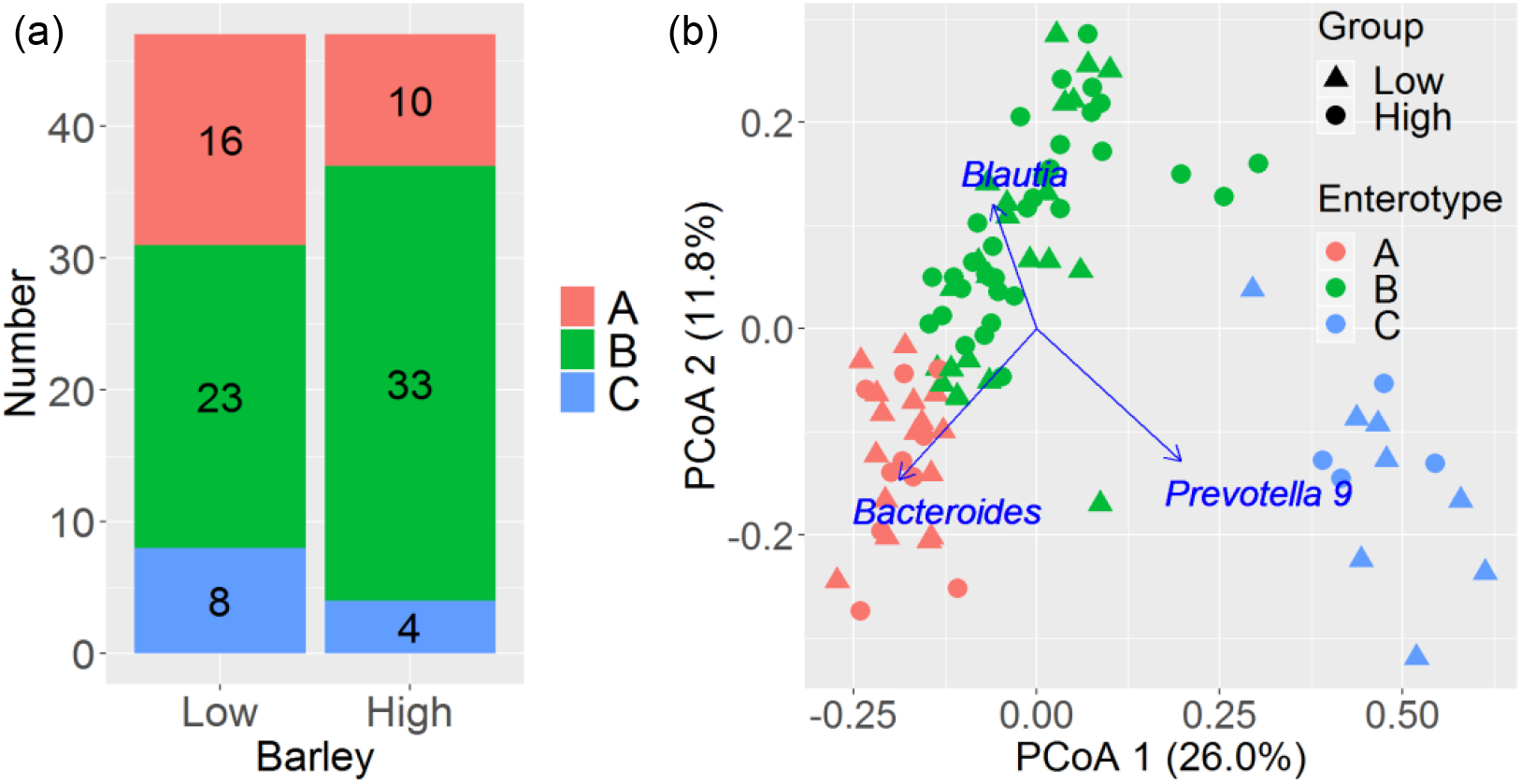
Microbiome enterotypes in high and low barley groups (n = 94) aged 19–65 years in 2018. (a) Comparison of numbers of enterotypes A, B, or C in each group. (b) Plot of PCoA. Colour indicates enterotype; symbols indicate high or low barley consumption. Arrows indicate top three environmental factors.

Clusters A and B were distributed in the negative direction of PCoA1 and cluster C in the positive direction. Clusters A and B were divided by PCoA2, and cluster B was in an intermediate position between A and C. The driven genera in each enterotype were *Bacteroides* (A), *Blautia* (B), and *Prevotella 9* (C) (correlation coefficients; all *P <* 0.001). The most common enterotypes of the 94 participants were in the following order: B (60%), A (28%), and C (13%). The distribution of enterotypes was similar between the barley groups (B > A > C) and tended to be in the positive direction of PCoA2 in the high barley group, but the values were not significant (correlation coefficient, *P =* 0.08, **Supplemental Figure 1**).

### Microbiome

Alpha-diversity did not significantly differ between the barley groups (**Supplemental Table 2**). The high barley group had a higher abundance of *Bifidobacterium* (*P =* 0.01), *Collinsella* (*P =* 0.03), *Anaerostipes* (*P =* 0.0502), *Butyricicoccus* (*P=*0.002), *Dialister* (*P =* 0.04), and *Ruminococcus 2* (*P =* 0.04) without FDR adjustment (**Table 3, Supplemental Table 3**). However, *Dialister* was not associated in the multiple regression analysis (barley dichotomy: *P* = 0.19, barley continuous: *P* = 0.92) and Kendall correlation (*r* = 0.08, *P* = 0.27) (**Supplemental Table 4**). Therefore, we excluded *Dialister* from the characteristic intestinal microbiota and the multiple regression analyses of all participants. Additionally, *Anaerostipes* was not significant (*P* = 0.0502) but appeared when using multiple regression (barley dichotomy; *P* = 0.0495; **Supplemental Table 4**). Therefore, we selected *Butyricicoccus, Bifidobacterium, Ruminococcus 2*, and *Anaerostipes* as candidate characteristic genera associated with barley consumption.

**Table 3.** Relative abundance (%) of microbiome bacterial genera in low and high barley groups

| Genus<br>(relative abundance %) | Low barley ($n = 47$)<br>0-31.8 g/1,000 kcal·d<sup>1</sup> | High barley ($n = 47$)<br>32.3-253 g/1,000 kcal·d<sup>1</sup> | $P^2$<br>(crude) | $P^3$<br>(FDR) |
| --- | --- | --- | --- | --- |
|  | Median<br>(interquartile range) | Median<br>(interquartile range) |  |  |
| *Bifidobacterium* | 2.73 (0.00, 5.10) | 5.61 (0.00, 8.12) | 0.01 | 0.37 |
| *Collinsella* | 0.92 (0.00, 1.82) | 1.95 (0.00, 2.20) | 0.03 | 0.42 |
| *Subdoligranulum* | 1.12 (0.00, 1.80) | 1.83 (0.00, 2.32) | 0.08 | 0.44 |
| *Anaerostipes* | 0.89 (0.02, 1.28) | 1.06 (0.07, 1.92) | 0.0502 | 0.42 |
| *Butyricicoccus* | 0.43 (0.00, 0.46) | 0.65 (0.12, 0.65) | 0.002 | 0.09 |
| Ruminococcaceae UCG-013 | 0.09 (0.00, 0.27) | 0.22 (0.00, 0.34) | 0.06 | 0.44 |
| *Dialister* | 0.00 (0.00, 0.31) | 0.07 (0.00, 0.43) | 0.04 | 0.42 |
| *Acidaminococcus* | 0.00 (0.00, 0.30) | 0.03 (0.00, 0.66) | 0.07 | 0.44 |
| *Ruminococcus 2* | 0.00 (0.00, 0.37) | 0.01 (0.00, 0.86) | 0.04 | 0.42 |

<sup>1</sup>Range of barley consumption rate
<sup>2</sup>Mann–Whitney $U$ -test (crude $P$ value)
<sup>3</sup>Mann–Whitney $U$ -test adjusted with FDR (False Discovery Rate) method

### Multiple regression analyses adjusted with dietary factors

Supplemental Table 5 shows that *Butyricicoccus* and *Ruminococcus 2* were associated with barley consumption after adjustment with dietary fibre, cereal, or legumes, but not with natto. Conversely, *Bifidobacterium* and *Anaerostipes* were not associated with barley consumption after adjustment with dietary fibre, cereal, legumes, and natto.

### Multiple regression analyses of all the participants

We assessed associations between barley and *Butyricicoccus, Bifidobacterium, Ruminococcus 2*, and *Anaerostipes* using multiple regression analyses (**Table 4**). *Butyricicoccus, Bifidobacterium*, and *Anaerostipes* were associated with barley intake, but *Ruminococcus 2* was not. Therefore, *Butyricicoccus, Bifidobacterium*, and *Anaerostipes* appeared to have a closer association with barley intake.

**Table 4.** Association between the relative abundance of microbiome bacteria and barley intake group, sex, age, and risk of diabetes, dyslipidemia, and hypertension by multivariate linear regression analyses (n = 236)

|  | *Butyricicoccus* | *Bifidobacterium* | *Ruminococcus 2* | *Anaerostipes* |
| --- | --- | --- | --- | --- |
| Independent variable (unit) | *R* (SE)<sup>1</sup> | *R* (SE)<sup>1</sup> | *R* (SE)<sup>1</sup> | *R* (SE)<sup>1</sup> |
| Barley group (0: low, 1: high) | 0.11 (0.05) * | 2.52 (1.00) * | 0.10 (0.15) | 0.79 (0.23) ** |
| Sex (0: female, 1: male) | 0.02 (0.06) | −1.31 (1.20) | −0.26 (0.19) | −0.31 (0.28) |
| Age (10 years) | 0.008 (0.024) | 0.36 (0.49) | 0.09 (0.07) | −0.09 (0.11) |
| Risk of diabetes (0–2) | 0.02 (0.07) | 0.45 (1.32) | 0.11 (0.20) | −0.26 (0.31) |
| Risk of dyslipidemia (0–2) | −0.02 (0.03) | −0.17 (0.60) | −0.0001 (0.09) | 0.13 (0.14) |
| Risk of hypertension (0–2) | −0.01 (0.04) | −1.31 (0.71)<sup>†</sup> | −0.11 (0.11) | −0.26 (0.11) |

Following characters mean significant differences. <sup>†</sup>: $P < 0.1$ , \*: $P < 0.05$ , \*\*: $P < 0.01$
<sup>1</sup> R: Linear regression coefficient, SE: standard error.

### Network analysis of microbiome

Supplemental Figure 2 shows the results of the network analysis. *Butyricicoccus* and *Bifidobacterium* were directly associated with barley intake and were classified into the same community group. *Anaerostipes* was not directly associated with barley intake but was associated via *Lachnospira*, Ruminococcaceae *UCG-13*, or *Tyzzerella 3* to barley intake. In addition, *Anaerostipes* belonged to the same community as barley, *Butyricicoccus*, and *Bifidobacterium. Ruminococcus 2* was directly associated with barley but did not belong to the same community group as barley, *Butyricicoccus, Bifidobacterium*, and *Anaerostipes*.

### Dietary habits as confounding factors

The results of the correlation analyses showed that dietary habits, consumption of fibre, and legumes were significantly associated, and polyunsaturated fatty acids tended to be associated with barley intake (Supplemental Table 6). Most healthy and all unhealthy dietary habits tended to correlate positively and negatively with barley intake, respectively.

## Discussion

We identified the characteristics of the gut microbiota in a Japanese population that consumes barley. Previous studies reported that the soluble fibre in barley increases the prevalence of intestinal bacteria, such as *Prevotella 9* and *Blautia*, and alpha-diversity ^(8, 4)^. In our study, *Bifidobacterium, Anaerostipes*, and *Butyricicoccus* abundance tended to increase (Table 4), and we consider these bacteria as specific candidates that relate to barley consumption. However, barley intake might have a limited effect because there were no changes in alpha-diversity.

The consumption of barley tended to be positively related to PCoA2, but this result was not significant (*p =* 0.08, **Supplemental Figure 1**). The numbers of enterotypes did not differ between the barley groups, and the absolute numbers of enterotype B were 33 (high) and 23 (low). These results suggested that barley consumption slightly shifted the enterotype to B; however, further studies are necessary to confirm this result. Arumugam *et al*. ^(9)^ reported a similar distribution in a “∧” shape (Figure 2b) that comprised enterotypes A (driven by *Bacteroides*), B (driven by *Blautia*), and C (driven by *Prevotella 9*). These results were consistent with those of a previous study; however, the influential microbiome of enterotype B, *Blautia*, was different ^(9)^. Arumugam *et al*. reported that the driven genera of enterotype B differed between cohorts, and they mentioned that *Blautia* was one of the driven genera^(9)^.

*Bifidobacterium, Anaerostipes*, and *Butyricicoccus* were identified as characteristic bacteria in the high barley group. *Bifidobacterium* was associated with the consumption of rye containing beta-glucan in a randomised control trial ^(20)^, *Butyricicoccus* was associated with barley in an animal study ^(21)^, but *Anaerostipes* was associated with omega-3 fatty acids ^(22)^. The present study found that the association between barley and *Anaerostipes* changed became non-significant after adjusting the analysis with other diet factors (Supplemental Table 5). Barley also tended to be associated with polyunsaturated fatty acids (Supplemental Table 6). These findings might explain why the network analysis directly associated barley with *Bifidobacterium* and *Butyricicoccus* but not with *Anaerostipes* (Supplemental Figure 2). Bacteria such as *Bifidobacterium, Anaerostipes*, and *Butyricicoccus* produce short-chain fatty acids (SCFAs) and contribute to host health. *Bifidobacterium* have been reported to produce SCFAs, such as acetic acid from carbohydrate ^(23)^, and are known as characteristic bacteria of Japanese people ^(1, 21)^. *Anaerostipes* is a butyric acid-producing bacteria involved in preventing colorectal cancer ^(24)^ and metabolic diseases ^(25)^. *Butyricicoccus* is a prevalent butyric acid producer associated with inflammatory bowel disease prevention ^(26)^. Additionally, SCFA production from dietary fibre leads to the acidification of the gut environment, which alters the gut microbiota composition ^(27)^. Therefore, these bacteria might have a positive influence on the hosts.

Furthermore, we identified some characteristic features of the Japanese cohort, for example, the relationships between barley and natto, as well as polyunsaturated fatty acids sourced from seafood. Thus, barley consumption is associated with the traditional Japanese diet.

This study has several limitations. The participants were employees of a company that manufactures barley products and would thus most likely consume more barley in general. The possibility of other confounding factors such as dietary habits cannot be completely excluded (Supplemental Tables 5, 6); thus, the effects of barley could be overestimated. We did not measure SCFAs or microbiome functions, which restricts the usefulness of our results. A follow-up intervention study with another cohort is warranted to overcome these limitations.

In conclusion, barley might change the intestinal microbiota of the Japanese population. We selected *Bifidobacterium, Anaerostipes*, and *Butyricicoccus* as candidate characteristic genera indicating barley consumption. It is important to consider that our findings might have been confounded by other dietary factors such as natto consumption.

## Data Availability

If requested, we will provide the data or will cooperate fully in obtaining and providing the data on which the manuscript is based for examination.

## Data Availability

All data used in this study were extracted from public GWAS summary statistics.

## Acknowledgments

### Source of funding, conflict of interests, authorship

This study was supported by funding from Hakubaku Co., Ltd. and National Institutes of Biomedical Innovation, Health and Nutrition. The department where Z.Y. works has received research grants for other studies from Hakubaku Co., Ltd. T.M., Y.G., M.N., S.N., and T.K. are employees of Hakubaku Co., Ltd. T.M., K.H., Y.G., H.M., T.K., H.Y., J.K. and Z.Y. Project administration: T.M., K.H., Y.G.,H.M., T.K., M.N., S.N. and K.K. Resources: H.M., K.K., M.M., K.H., J.P., H.K., K.M., and J.K.Investigation and Writing original draft: T.M., K.H. and J.P. Writing, revising and editing: T.M., J.K. and Z.Y. All authors have read and approved the final manuscript.

## Transparency Declaration

The lead author affirms that this manuscript is an honest, accurate, and transparent account of the study being reported. The reporting of this work is compliant with STROBE guidelines. The lead author affirms that no important aspects of the study have been omitted and that any discrepancies from the study as planned have been explained.

## Supporting Information

Supplemental Tables 1–6 and Supplemental Figures 1–2 are available from the Online Supplemental Material. Data described in the manuscript, code book, and analytic code will be made available after approval of our request.

## References

1. Nishijima S, Suda W, Oshima K et al. (2016) The gut microbiome of healthy Japanese and its microbial and functional uniqueness. DNA Res 23, 125–33. doi: 10.1093/dnares/dsw002

2. Sonnenburg ED & Sonnenburg JL (2014) Starving our microbial self: The deleterious consequences of a diet deficient in microbiota-accessible carbohydrates. Cell Metab. 20 779–86. doi: https://doi.org/10.1016/j.cmet.2014.07.003

3. Matsuoka T, Tsuchida A, Yamaji A et al. (2020) Consumption of a meal containing refined barley flour bread is associated with a lower postprandial blood glucose concentration after a second meal compared with one containing refined wheat flour bread in healthy Japanese: A randomized control trial. Nutrition 72, 110637. doi: 10.1016/j.nut.2019.110637

4. Martínez I, Lattimer JM, Hubach KL et al. (2013) Gut microbiome composition is linked to whole grain-induced immunological improvements. ISME J. 7, 269–80. doi: https://doi.org/10.1038/ismej.2012.104

5. Kobayashi T, Kaneko S, Matsuoka T (2013) The effect of barley noodles on blood sugar levels in type 2 diabetes patients. J Japanese Assoc Diet Fiber Res 17 35–40. Available from: https://ci.nii.ac.jp/naid/40019769176

6. Matsuoka T, Uchimatsu D, Kobayashi T et al. (2014) Effect of barley on metabolic syndrome related indicators in overweight Japanese men and women. J Japanese Assoc Diet Fiber Res 18 25–33. Available from: https://ci.nii.ac.jp/naid/40020187747

7. Nilsson AC, Östman EM, Knudsen KEB et al. (2010) A cereal-based evening meal rich in indigestible carbohydrates increases plasma butyrate the next morning. J Nutr 140 1932–6. doi: 10.3945/jn.110.123604

8. Kovatcheva-Datchary P, Nilsson A, Akrami R et al. (2015) Dietary fiber-induced improvement in glucose metabolism is associated with increased abundance of Prevotella. Cell Metab 22 971–82. doi: 10.1016/j.cmet.2015.10.001

9. Arumugam M, Raes J, Pelletier E et al. (2011) Enterotypes of the human gut microbiome. Nature 473 174–80. doi: 10.1038/nature09944i

10. Nilsson AC, Ostman EM, Granfeldt Y et al. (2008) Effect of cereal test breakfasts differing in glycemic index and content of indigestible carbohydrates on daylong glucose tolerance in healthy subjects. Am J Clin Nutr 87 645–54. doi: 10.1093/ajcn/87.3.645

11. Lifschitz CH, Grusak MA, Butte NF (2002) Carbohydrate digestion in humans from a beta-glucan-enriched barley is reduced. J Nutr 132 2593–6. doi: 10.1093/jn/132.9.2593

12. Hosomi K, Ohno H, Murakami H et al. (2017) Method for preparing DNA from feces in guanidine thiocyanate solution affects 16S rRNA-based profiling of human microbiota diversity. Sci Rep 7 4339. doi: 10.1038/s41598-017-04511-0

13. Caporaso JG, Kuczynski J, Stombaugh J et al. (2010) QIIME allows analysis of high-throughput community sequencing data. Nat Methods 7 335–6. doi: 10.1038/nmeth.f.303

14. Mohsen A, Park J, Chen Y-A et al. (2019) Impact of quality trimming on the efficiency of reads joining and diversity analysis of Illumina paired-end reads in the context of QIIME1 and QIIME2 microbiome analysis frameworks. BMC Bioinformatics 20 581. doi: 10.1186/s12859-019-3187-5

15. Edgar RC (2010) Search and clustering orders of magnitude faster than BLAST. Bioinformatics 26 2460–1. doi: 10.1093/bioinformatics/btq461

16. Quast C, Pruesse E, Yilmaz P et al. (2013) The SILVA ribosomal RNA gene database project: improved data processing and web-based tools. Nucleic Acids Res 41 D590–6. doi: 10.1093/nar/gks1219

17. GBD 2017 Diet Collaborators (2019) Health effects of dietary risks in 195 countries, 1990-2017: a systematic analysis for the Global Burden of Disease Study 2017. Lancet 393 1958–72. doi: 10.1016/S0140-6736(19)30041-8

18. Ito K, Murphy D (2013) Tutorial: Application of ggplot2 to pharmacometric graphics. CPT Pharmacometrics Syst Pharmacol 2 1–16. doi: 10.1038/psp.2013.56

19. Casella G, Fienberg S, Olkin I (2006) A Modern Approach to Regression with R. Vol. 102, Design. 618 p.

20. Eriksen AK, Brunius C, Mazidi M et al. (2020) Effects of whole-grain wheat, rye, and lignan supplementation on cardiometabolic risk factors in men with metabolic syndrome: a randomized crossover trial. Am J Clin Nutr 111, 864–76. doi: 10.1093/ajcn/nqaa026

21. Moen B, Berget I, Rud I et al. (2016) Extrusion of barley and oat influence the fecal microbiota and SCFA profile of growing pigs. Food Funct 7 1024–32. doi: 10.1039/c5fo01452b

22. Noriega BS, Sanchez-Gonzalez MA, Salyakina D et al. (2016) Understanding the impact of Omega-3 rich diet on the gut microbiota. Case Rep Med, 3089303. doi: 10.1155/2016/3089303

23. Pratt C, Campbell MD (2020) The effect of bifidobacterium on reducing symptomatic abdominal pain in patients with irritable bowel syndrome: A systematic review. Probiotics Antimicrob Proteins 12 834–9. doi: 10.1007/s12602-019-09609-7

24. Ai D, Pan H, Li X et al. (2019) Identifying gut microbiota associated with colorectal cancer using a zero-inflated lognormal model. Front Microbiol 10, 826. doi: 10.3389/fmicb.2019.00826

25. Zeevi D, Korem T, Godneva A et al. (2019) Structural variation in the gut microbiome associates with host health. Nature 568 43–8. doi: 10.1038/s41586-019-1065-y

26. Eeckhaut V, Machiels K, Perrier C et al. (2013) Butyricicoccus pullicaecorum in inflammatory bowel disease. Gut 62,1745–52. doi: 10.1136/gutjnl-2012-303611

27. Scott KP, Duncan SH, Flint HJ (2008) Dietary fibre and the gut microbiota. Nutr Bull 33 201–11. Available from: http://doi.wiley.com/10.1111/j.1467-3010.2008.00706.x

