## Supplemental Table, Supplemental Figure for "Relationships between barley consumption and gut microbiome characteristics in a healthy Japanese population: a cross-sectional study"

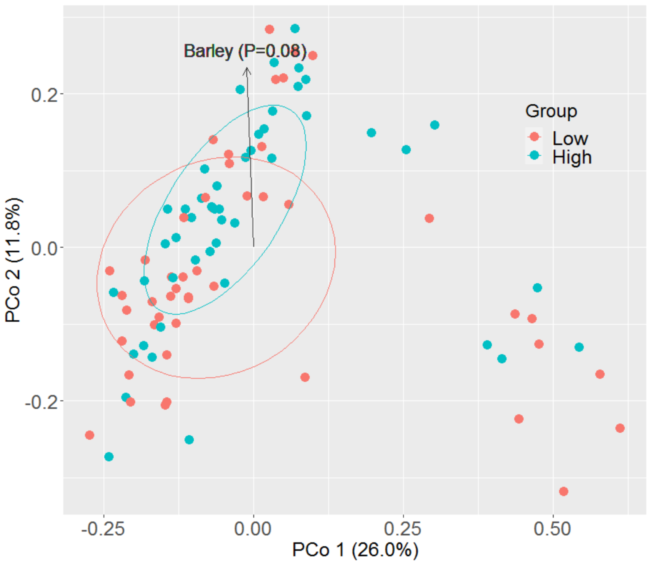


**Supplemental Figure 1** PCoA result. Color means high or low consumption of barley. The arrow means high barley group (*n* 94, aged 19-65 years, Japan, 2018).

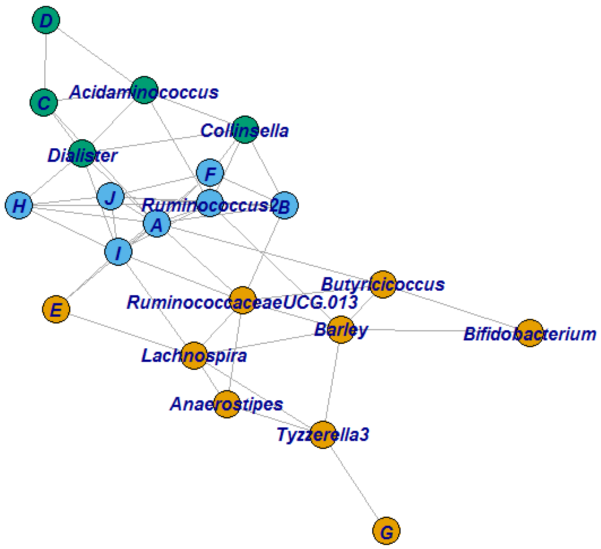


**Supplemental Figure 2** The result of network analysis. Line means correlation |*r*|>0.15 (Kendall rank-sum tests). Colors mean community groups. Described names are those in which the difference between low barley and high barley is *P*<0.1 (Mann-Whitney *U*-test) (*n* 94, aged 19-65 years, Japan, 2018).

The bacteria described as characters are as follow; *A*: *Subdoligranulum*, *B*: *Eubacterium Hallii group*, *C*: *Prevotella2*, *D*: *Megasphaera*, *E*: *Roseburia*, *F*: *Veillonella*, *G*: *Lachnospiraceae family uncultured*, *H*: *Dorea*, *I*: *Ruminococcus1*, *J*: *Eubacterium coprostanoligenes group*.

**Supplemental Table 1** Criterion of risk group of diabetes, hypertension, and dislipidemia.

| Variable | *n* (Male) | Low barley (*n* 119) | High barley (*n* 117) | Criterion |
|---|---|---|---|---|
| Diabetes risk | | | | |
| Risk 2 | 8 (8) | 3 | 5 | HbA1c>=6.5% or Fasting blood glucose>=126 mg/dL or Under medication |
| Risk 1 | 5 (5) | 4 | 1 | Who were not Risk 2, and Fasting blood glucose>=110 mg/dL |
| Risk 0 | 223 (161) | 110 | 113 | Who were neither Risk 1 nor Risk 2 |
| Hypertension risk | | | | |
| Risk 2 | 46 (43) | 20 | 26 | SBP>=140 mmHg or DBP>=90 mmHg or Under medication |
| Risk 1 | 20 (16) | 11 | 9 | Who were not Risk 2, and either SBP>=130 mgHg or DBP>=85 mgHg |
| Risk 0 | 170 (115) | 86 | 84 | Who were neither Risk 1 nor Risk 2 |
| Dislipidemia risk | | | | |
| Risk 2 | 97 (84) | 46 | 51 | TG>=150 mg/dL or HDL-cholesterol<40 mg/dL or LDL-cholesterol>=140 mg/dL or Under medication |
| Risk 1 | 31 (25) | 17 | 14 | Who were not Risk 2, and LDL-cholesterol>=120 mg/dL |
| Risk 0 | 108 (65) | 54 | 54 | Who were neither Risk 1 nor Risk 2 |

Risk 2 means disease, Risk 1 means border line, and Risk 0 means healthy participants.

Range of barley consumption; Low barley: 0-31.8 g/1000 kcal･day, High barley: 32.3-253 g/1000 kcal･day

**Supplemental Table 2** Alpha-diversity, compared low barley with high barley group on cross-sectional study (aged 19-65 years in 2018, Japan).

| | Low barley (*n* 47) range: 0-31.8 g/1000 kcal･d | High barley (*n* 47) range: 32.3-253 g/1000 kcal･d | | |
|---|---|---|---|---|
| Variable | Median [ Interquartile range ] | Median [ Interquartile range ] | *P* value[1)] | $P_{FDR}$ value[2)] |
| Observed | 561 [ 482, 649 ] | 549 [ 500, 628 ] | 0.86 | 0.87 |
| Chao1 | 1146 [ 919, 1290 ] | 1170 [ 971, 1419 ] | 0.41 | 0.68 |
| Shannon | 3.77 [ 3.46, 3.95 ] | 3.77 [ 3.57, 4.14 ] | 0.38 | 0.68 |
| Simpson | 0.94 [ 0.90, 0.95 ] | 0.94 [ 0.92, 0.96 ] | 0.21 | 0.68 |
| Fisher | 128 [ 106, 156 ] | 125 [ 111, 149 ] | 0.87 | 0.87 |

[1)] Compared low and high barley groups using Mann–Whitney *U* -test (crude *P* value)

[2)] Compared low and high barley groups using Mann–Whitney *U* -test adjusted with *FDR* method

**Supplemental Table 3** Relative abundance(%) of the frequent 50 microbiomes (genus levels), compared low barley with high barley group on cross-sectional study (aged 19-65 years in 2018, Japan).

| | Low barley (*n* 47) range: 0-31.8 g/1000 kcal･d | High barley (*n* 47) range: 32.3-253 g/1000 kcal･d | | |
|---|---|---|---|---|
| Genus | Median [ Interquartile range ] | Median [ Interquartile range ] | *P* value[1)] | $P_{FDR}$ value[2)] |
| *Bacteroides* | 31.27 [ 0.01, 28.98 ] | 29.28 [ 1.71, 27.52 ] | 0.50 | 0.79 |
| *Blautia* | 6.15 [ 0.78, 7.48 ] | 5.81 [ 1.63, 6.67 ] | 0.64 | 0.85 |
| *Bifidobacterium* | 2.73 [ 0.00, 5.10 ] | 5.61 [ 0.00, 8.12 ] | 0.01 | 0.37 |
| *Faecalibacterium* | 5.76 [ 0.00, 6.02 ] | 6.08 [ 0.00, 6.34 ] | 0.54 | 0.79 |
| *Prevotella 9* | 0.01 [ 0.00, 6.96 ] | 0.00 [ 0.00, 3.90 ] | 0.20 | 0.59 |
| *Eubacterium rectale group* | 1.10 [ 0.00, 2.28 ] | 1.56 [ 0.00, 2.62 ] | 0.73 | 0.87 |
| *Parabacteroides* | 2.28 [ 0.00, 2.62 ] | 1.64 [ 0.00, 1.96 ] | 0.18 | 0.59 |
| *Subdoligranulum* | 1.12 [ 0.00, 1.80 ] | 1.83 [ 0.00, 2.32 ] | 0.08 | 0.44 |
| *Collinsella* | 0.92 [ 0.00, 1.82 ] | 1.95 [ 0.00, 2.20 ] | 0.03 | 0.42 |
| *Sutterella* | 1.41 [ 0.00, 1.82 ] | 1.43 [ 0.00, 2.02 ] | 0.99 | 0.99 |
| *Megamonas* | 0.00 [ 0.00, 3.03 ] | 0.00 [ 0.00, 0.76 ] | 0.20 | 0.59 |
| *Ruminococcus torques group* | 1.27 [ 0.03, 1.67 ] | 1.09 [ 0.01, 1.73 ] | 0.36 | 0.72 |
| *Anaerostipes* | 0.89 [ 0.02, 1.28 ] | 1.06 [ 0.07, 1.92 ] | 0.0502 | 0.42 |
| *Lachnoclostridium* | 1.22 [ 0.19, 1.69 ] | 1.23 [ 0.04, 1.48 ] | 0.53 | 0.79 |
| *Fusicatenibacter* | 1.41 [ 0.00, 1.49 ] | 1.00 [ 0.00, 1.40 ] | 0.83 | 0.90 |
| *Fusobacterium* | 0.00 [ 0.00, 1.14 ] | 0.00 [ 0.00, 1.65 ] | 0.91 | 0.95 |
| *Eubacterium hallii group* | 0.97 [ 0.00, 1.28 ] | 0.78 [ 0.00, 0.97 ] | 0.60 | 0.83 |
| *Alistipes* | 0.21 [ 0.00, 0.82 ] | 0.40 [ 0.00, 1.30 ] | 0.34 | 0.72 |
| *Lachnospira* | 0.41 [ 0.00, 0.78 ] | 0.62 [ 0.00, 1.10 ] | 0.22 | 0.59 |

| | | | | |
|---|---|---|---|---|
| *Prevotella 2* | 0.00 [ 0.00, 0.34 ] | 0.00 [ 0.00, 1.46 ] | 0.99 | 0.99 |
| *Megasphaera* | 0.00 [ 0.00, 0.65 ] | 0.00 [ 0.00, 1.15 ] | 0.28 | 0.66 |
| *Roseburia* | 0.66 [ 0.00, 1.04 ] | 0.40 [ 0.00, 0.72 ] | 0.18 | 0.59 |
| *Veillonella* | 0.03 [ 0.00, 1.06 ] | 0.03 [ 0.00, 0.70 ] | 0.47 | 0.79 |
| *Phascolarctobacterium* | 0.01 [ 0.00, 0.79 ] | 0.00 [ 0.00, 0.70 ] | 0.58 | 0.83 |
| *Escherichia Shigella* | 0.07 [ 0.00, 0.72 ] | 0.05 [ 0.00, 0.75 ] | 0.73 | 0.87 |
| *Lachnospiraceae family uncultured* | 0.49 [ 0.01, 0.72 ] | 0.59 [ 0.00, 0.70 ] | 0.91 | 0.95 |
| *Alloprevotella* | 0.00 [ 0.00, 0.56 ] | 0.00 [ 0.00, 0.74 ] | 0.77 | 0.89 |
| *Dorea* | 0.40 [ 0.00, 0.81 ] | 0.35 [ 0.00, 0.43 ] | 0.32 | 0.72 |
| *Ruminococcus 2* | 0.00 [ 0.00, 0.37 ] | 0.01 [ 0.00, 0.86 ] | 0.04 | 0.42 |
| *Butyricicoccus* | 0.43 [ 0.00, 0.46 ] | 0.65 [ 0.12, 0.65 ] | 0.00 | 0.09 |
| *Lachnospiraceae* UCG-008 | 0.52 [ 0.00, 0.57 ] | 0.32 [ 0.00, 0.48 ] | 0.81 | 0.90 |
| *Ruminococcus 1* | 0.01 [ 0.00, 0.28 ] | 0.01 [ 0.00, 0.69 ] | 0.34 | 0.72 |
| *Acidaminococcus* | 0.00 [ 0.00, 0.30 ] | 0.03 [ 0.00, 0.66 ] | 0.07 | 0.44 |
| *Ruminococcaceae family uncultured* | 0.23 [ 0.00, 0.42 ] | 0.24 [ 0.00, 0.54 ] | 0.64 | 0.85 |
| *Parasutterella* | 0.02 [ 0.00, 0.38 ] | 0.02 [ 0.00, 0.57 ] | 0.50 | 0.79 |
| *Streptococcus* | 0.12 [ 0.00, 0.52 ] | 0.20 [ 0.00, 0.43 ] | 0.50 | 0.79 |
| *Mitsuokella* | 0.00 [ 0.00, 0.59 ] | 0.00 [ 0.00, 0.35 ] | 0.38 | 0.73 |
| *Prevotellaceae NK3B31 group* | 0.00 [ 0.00, 0.48 ] | 0.00 [ 0.00, 0.28 ] | 0.71 | 0.87 |
| *Dialister* | 0.00 [ 0.00, 0.31 ] | 0.07 [ 0.00, 0.43 ] | 0.04 | 0.42 |
| *Ruminiclostridium 5* | 0.12 [ 0.00, 0.36 ] | 0.15 [ 0.01, 0.32 ] | 0.66 | 0.85 |
| *Ruminococcaceae* UCG-013 | 0.09 [ 0.00, 0.27 ] | 0.22 [ 0.00, 0.34 ] | 0.06 | 0.44 |
| *Barnesiella* | 0.00 [ 0.00, 0.23 ] | 0.05 [ 0.00, 0.36 ] | 0.22 | 0.59 |
| *Ruminococcaceae* UCG-002 | 0.00 [ 0.00, 0.17 ] | 0.01 [ 0.00, 0.39 ] | 0.19 | 0.59 |

| | | | | |
|---|---|---|---|---|
| *Rhodospirillaceae family uncultured* | 0.00 [ 0.00, 0.26 ] | 0.00 [ 0.00, 0.27 ] | 0.43 | 0.79 |
| *Ruminococcus gauvreauii group* | 0.00 [ 0.00, 0.23 ] | 0.00 [ 0.00, 0.25 ] | 0.79 | 0.90 |
| *Paraprevotella* | 0.00 [ 0.00, 0.31 ] | 0.00 [ 0.00, 0.17 ] | 0.20 | 0.59 |
| *Odoribacter* | 0.04 [ 0.00, 0.21 ] | 0.10 [ 0.00, 0.24 ] | 0.48 | 0.79 |
| *Eubacterium coprostanoligenes group* | 0.00 [ 0.00, 0.14 ] | 0.01 [ 0.00, 0.30 ] | 0.21 | 0.59 |
| *Bilophila* | 0.10 [ 0.00, 0.18 ] | 0.14 [ 0.00, 0.25 ] | 0.24 | 0.60 |
| *Tyzzerella 3* | 0.00 [ 0.00, 0.10 ] | 0.00 [ 0.00, 0.29 ] | 0.22 | 0.59 |

1) Compared low and high barley groups using Mann–Whitney *U* -test (crude *P* value)

2) Compared low and high barley groups using Mann–Whitney *U* -test adjusted with *FDR* method

**Supplemental Table 4** Results of Multiple regression analysis (dichotomy and continuous) and Kendall rank correlation between barley consumption rate (g/1000 kcal･day) and intestinal microbiota on cross-sectional study (aged 19-65 years in 2018, Japan).

| | Multiple regression (Barley:dichotomy)[1)] | | | Multiple regression (Barley:continuous)[2)] | | | Kendall rank correlation[3)] | |
|---|---|---|---|---|---|---|---|---|
| Genus | Estimate | SE | *P* value | Estimate | SE | *P* value | Estimate | *P* value |
| *Bacteroides* | -1.51 | 3.15 | 0.63 | -0.06 | 0.04 | 0.57 | -0.05 | 0.48 |
| *Blautia* | -0.85 | 0.85 | 0.32 | -0.02 | 0.01 | 0.50 | -0.05 | 0.47 |
| *Bifidobacterium* | 2.78 | 1.63 | 0.09 | 0.11 | 0.02 | 0.04 | 0.17 | 0.01 |
| *Faecalibacterium* | -0.21 | 1.09 | 0.85 | -0.04 | 0.01 | 0.23 | 0.07 | 0.32 |
| *Prevotella 9* | -2.60 | 2.91 | 0.38 | -0.07 | 0.03 | 0.44 | -0.09 | 0.27 |
| *Eubacterium rectale group* | 0.55 | 0.58 | 0.34 | 0.02 | 0.01 | 0.37 | 0.02 | 0.80 |
| *Parabacteroides* | -0.65 | 0.39 | 0.10 | -0.01 | 0.00 | 0.35 | -0.11 | 0.12 |
| *Subdoligranulum* | 0.43 | 0.46 | 0.36 | -0.01 | 0.01 | 0.57 | 0.06 | 0.38 |
| *Collinsella* | 0.50 | 0.52 | 0.34 | 0.02 | 0.01 | 0.15 | 0.12 | 0.10 |
| *Sutterella* | 0.24 | 0.44 | 0.59 | -0.01 | 0.01 | 0.42 | -0.03 | 0.67 |
| *Megamonas* | -1.87 | 1.15 | 0.11 | -0.06 | 0.01 | 0.13 | -0.11 | 0.19 |
| *Ruminococcus torques group* | 0.16 | 0.41 | 0.70 | 0.00 | 0.00 | 0.94 | -0.05 | 0.45 |
| *Anaerostipes* | 0.71 | 0.36 | 0.05 | 0.01 | 0.00 | 0.36 | 0.08 | 0.25 |
| *Lachnoclostridium* | -0.17 | 0.27 | 0.54 | 0.00 | 0.00 | 0.96 | -0.04 | 0.54 |
| *Fusicatenibacter* | -0.06 | 0.27 | 0.82 | -0.01 | 0.00 | 0.46 | -0.02 | 0.80 |
| *Fusobacterium* | 0.74 | 0.85 | 0.39 | 0.01 | 0.01 | 0.61 | -0.03 | 0.72 |
| *Eubacterium hallii group* | -0.42 | 0.24 | 0.08 | -0.01 | 0.00 | 0.50 | 0.00 | 0.98 |
| *Alistipes* | 0.29 | 0.37 | 0.44 | 0.00 | 0.00 | 1.00 | 0.05 | 0.44 |
| *Lachnospira* | 0.23 | 0.25 | 0.36 | 0.01 | 0.00 | 0.26 | 0.16 | 0.03 |
| *Prevotella 2* | 1.41 | 0.64 | 0.03 | 0.01 | 0.01 | 0.68 | -0.01 | 0.90 |
| *Megasphaera* | 0.74 | 0.60 | 0.22 | 0.08 | 0.01 | 0.00 | 0.06 | 0.43 |
| *Roseburia* | -0.40 | 0.22 | 0.08 | -0.01 | 0.00 | 0.16 | -0.07 | 0.32 |
| *Veillonella* | -0.37 | 0.44 | 0.40 | 0.00 | 0.01 | 0.86 | 0.14 | 0.07 |

| | | | | | | | | |
|---|---|---|---|---|---|---|---|---|
| *Phascolarctobacterium* | -0.19 | 0.22 | 0.39 | 0.00 | 0.00 | 0.66 | 0.00 | 0.96 |
| *Escherichia Shigella* | 0.06 | 0.47 | 0.90 | 0.02 | 0.01 | 0.26 | 0.03 | 0.69 |
| *Lachnospiraceae family uncultured* | 0.02 | 0.14 | 0.90 | 0.01 | 0.00 | 0.01 | 0.07 | 0.31 |
| *Alloprevotella* | 0.18 | 0.56 | 0.75 | -0.01 | 0.01 | 0.56 | -0.06 | 0.49 |
| *Dorea* | -0.32 | 0.19 | 0.10 | -0.01 | 0.00 | 0.39 | -0.06 | 0.36 |
| *Ruminococcus 2* | 0.45 | 0.23 | 0.06 | 0.00 | 0.00 | 0.51 | 0.15 | 0.05 |
| *Butyricicoccus* | 0.17 | 0.07 | 0.02 | 0.00 | 0.00 | 0.05 | 0.21 | 0.00 |
| *Lachnospiraceae* UCG-008 | -0.11 | 0.11 | 0.32 | 0.00 | 0.00 | 0.60 | 0.01 | 0.93 |
| *Ruminococcus 1* | 0.36 | 0.21 | 0.08 | 0.01 | 0.00 | 0.18 | 0.10 | 0.20 |
| *Acidaminococcus* | 0.37 | 0.20 | 0.07 | 0.00 | 0.00 | 0.82 | 0.08 | 0.29 |
| *Ruminococcaceae family uncultured* | 0.03 | 0.13 | 0.81 | 0.01 | 0.00 | 0.15 | 0.01 | 0.85 |
| *Parasutterella* | 0.18 | 0.20 | 0.38 | 0.02 | 0.00 | 0.01 | 0.08 | 0.27 |
| *Streptococcus* | -0.11 | 0.17 | 0.53 | 0.00 | 0.00 | 0.79 | 0.07 | 0.32 |
| *Mitsuokella* | -0.21 | 0.45 | 0.65 | 0.00 | 0.01 | 0.93 | 0.02 | 0.84 |
| *Prevotellaceae NK3B31 group* | -0.34 | 0.41 | 0.41 | -0.01 | 0.00 | 0.53 | 0.00 | 0.99 |
| *Dialister* | 0.17 | 0.13 | 0.19 | 0.00 | 0.00 | 0.92 | 0.08 | 0.27 |
| *Ruminiclostridium 5* | -0.04 | 0.11 | 0.72 | 0.00 | 0.00 | 0.95 | 0.01 | 0.89 |
| *Ruminococcaceae* UCG-013 | 0.06 | 0.08 | 0.49 | 0.00 | 0.00 | 0.27 | 0.17 | 0.02 |
| *Barnesiella* | 0.09 | 0.12 | 0.46 | 0.00 | 0.00 | 0.63 | 0.08 | 0.29 |
| *Ruminococcaceae* UCG-002 | 0.14 | 0.13 | 0.25 | 0.00 | 0.00 | 0.98 | 0.07 | 0.38 |
| *Rhodospirillaceae family uncultured* | 0.02 | 0.21 | 0.92 | 0.00 | 0.00 | 0.64 | -0.09 | 0.25 |
| *Ruminococcus gauvreauii group* | 0.01 | 0.11 | 0.93 | 0.00 | 0.00 | 0.38 | -0.01 | 0.93 |
| *Paraprevotella* | -0.15 | 0.13 | 0.23 | 0.00 | 0.00 | 0.86 | -0.13 | 0.11 |
| *Odoribacter* | 0.01 | 0.08 | 0.87 | 0.00 | 0.00 | 0.88 | 0.02 | 0.76 |
| *Eubacterium coprostanoligenes group* | 0.13 | 0.10 | 0.21 | 0.01 | 0.00 | 0.03 | 0.07 | 0.38 |
| *Bilophila* | 0.05 | 0.05 | 0.29 | 0.00 | 0.00 | 0.46 | 0.06 | 0.40 |
| *Tyzzerella 3* | 0.19 | 0.13 | 0.16 | 0.02 | 0.00 | 0.00 | 0.16 | 0.04 |

[1)] Results of multiple regression analysis. Genus levels of microbiome relative abundance (%) were used as the dependent variable, barley consumption rate (0="Low barley group", 1="High barley group"), age (years), and sex (0="female", 1="male") were used as the independent variable. Estimate: Linear regression coefficient, SE: standard error.

[2)] Results of multiple regression analysis. Genus levels of microbiome relative abundance (%) were used as the dependent variable, barley consumption rate (g/1000 kcal･day), age (years), and sex (0="female", 1="male") were used as the independent variable. Estimate: Linear regression coefficient, SE: standard error.

[3)] Results of correlation test between barley consumption rate (g/1000 kcal･day) and genus levels of intestinal microbiota relative abundance (%) using Kendall method. Estimate: Kendall's correlation coefficient.

**Supplemental Table 5** Cross-sectional relationship between barley intake group (Low = 0, High = 1) and microbiomes (*Butyricicoccus* , *Bifidobacterium* , *Ruminococcus 2* , and *Anaerostipes* ) using multiple regression model (aged 19-65 years in 2018, Japan).

| | Dependent variable | | | |
|---|---|---|---|---|
| Independent variable (unit) | *Butyricicoccus* | *Bifidobacterium* | *Ruminococcus 2* | *Anaerostipes* |
| Model 1 | | | | |
| Barley group (Low = 0, High = 1) | 0.165 (0.077) * | 3.253 (1.727) † | 0.527 (0.246) * | 0.594 (0.377) |
| Sex (Female =0, Male = 1) | -0.061 (0.075) | -1.238 (1.691) | -0.23 (0.241) | 0.328 (0.369) |
| Age (10 years) | -0.026 (0.034) | 0.909 (0.768) | 0.011 (0.109) | -0.153 (0.167) |
| Dietary fiber (g/1000 kcal) | 0.008 (0.024) | -0.454 (0.546) | -0.073 (0.078) | 0.110 (0.119) |
| Model 2 | | | | |
| Barley group (Low = 0, High = 1) | 0.173 (0.075) * | 2.321 (1.682) | 0.601 (0.233) * | 0.645 (0.369) † |
| Sex (Female =0, Male = 1) | -0.066 (0.074) | -1.200 (1.664) | -0.095 (0.231) | 0.218 (0.365) |
| Age (10 years) | -0.025 (0.036) | 1.174 (0.819) | -0.105 (0.113) | -0.094 (0.179) |
| Cereal (g/1000 kcal) | 0.000 (0.001) | 0.017 (0.015) | -0.005 (0.002) * | 0.002 (0.003) |
| Model 3 | | | | |
| Barley group (Low = 0, High = 1) | 0.165 (0.073) * | 2.621 (1.663) | 0.503 (0.236) * | 0.675 (0.363) † |
| Sex (Female =0, Male = 1) | -0.063 (0.073) | -0.880 (1.657) | -0.198 (0.235) | 0.265 (0.362) |
| Age (10 years) | -0.029 (0.035) | 0.764 (0.781) | 0.028 (0.111) | -0.155 (0.171) |
| Legumes (g/1000 kcal) | 0.001 (0.002) | 0.025 (0.046) | -0.008 (0.006) | 0.005 (0.010) |
| Model 4 | | | | |
| Barley group (Low = 0, High = 1) | 0.150 (0.077) † | 2.259 (1.732) | 0.487 (0.248) † | 0.712 (0.380) † |
| Sex (Female =0, Male = 1) | -0.060 (0.073) | -0.809 (1.656) | -0.189 (0.237) | 0.253 (0.363) |
| Age (10 years) | -0.030 (0.034) | 0.723 (0.776) | 0.009 (0.111) | -0.138 (0.170) |
| *Natto* (g/1000 kcal) | 0.004 (0.004) | 0.082 (0.091) | -0.006 (0.013) | -0.001 (0.020) |

Following characters mean significant differences. †:$P<0.1$, *:$P<0.05$

Results of multiple regression analysis. Genus levels of microbiome relative abundance (%) were used as the dependent variable.

Barley consumption rate (0="Low barley group", 1="High barley group"), age (years), sex (0="female", 1="male"), and dietary factor (dietary fiber, cereal, legumes, *natto* , g/1000 kcal) were used as the independent variable.

Data are Linear regression coefficient and standard error.

**Supplemental Table 6** Correlation test between barley intake and dietary custom on cross-sectional study (aged 19-65 years in 2018, Japan).

| Variable | Estimate | *P* value |
|---|---|---|
| Healthy diet and nutrients | | |
| Dietary fibre (g/1000 kcal･d) | 0.31 | <0.001 |
| Legumes (g/1000 kcal･d) | 0.15 | 0.03 |
| Polyunsaturated fatty acids (g/1000 kcal･d) | 0.14 | 0.051 |
| Omega-3 fatty acids (g/1000 kcal･d) | 0.10 | 0.15 |
| Green vegetables (g/1000 kcal･d) | 0.07 | 0.32 |
| Calcium (mg/1000 kcal･d) | 0.06 | 0.41 |
| Fruits (g/1000 kcal･d) | -0.01 | 0.92 |
| Milk (g/1000 kcal･d) | -0.06 | 0.40 |
| Unhealthy diet and nutrients | | |
| Red meat (g/1000 kcal･d) | -0.01 | 0.94 |
| Sodium (mg/1000 kcal･d) | -0.07 | 0.35 |
| Sugar-sweetened beverages (g/1000 kcal･d) | -0.10 | 0.14 |

Kendall rank correlation between barley intake (kcal/1000 kcal･d) and diet or nutrients (weight /1000 kcal･d)

Vegetables: green vegetables

Milk: include yoghurt
